# Secondary Prevention of Cardiovascular Events in Patients with Overweight/Obesity in Routine Clinical Practice

**DOI:** 10.64898/2026.02.18.26346594

**Authors:** Wenxin Guo, Maidou Wang, Jiwon Shin, Fan Li, Emily C. O’Brien, LáShauntá Glover, Kristina Bortfeld, Anqi Zhao, Ryan McDevitt, Cheryl Kalapura, Sarah Wu, Sahar Shibeika, Shannon Aymes, Michael Porter, Brian Mac Grory, Jay B. Lusk

## Abstract

**Background and Aims:** The glucagon-like peptide-1 receptor agonist (GLP-1 RA) semaglutide has demonstrated efficacy for the secondary prevention of cardiovascular disease among patients with overweight/obesity without diabetes mellitus. However, the comparative effectiveness of GLP-1 RA versus other antiobesity medications (e.g. phentermine-topiramate) not been evaluated.

**Methods:** This was a retrospective, observational, cohort study using target trial emulation methodology using the Truveta electronic health record database of more than 120 million patients. Adult patients with a body mass index (BMI) >=27 kg/m^2^, a history of cardiovascular disease (prior ischemic stroke, transient ischemic attack, or myocardial infarction, or known coronary artery disease, heart failure, or peripheral artery disease) without diabetes mellitus were included in the study. The primary endpoint was time to first major adverse cardiovascular or cerebrovascular event (MACCE, defined as stroke or myocardial infarction).

**Results:** In total, 35,240 were included in the bupropion-naltrexone versus GLP-1 RA comparison, and 27,051 were included in the phentermine-topiramate versus GLP-1 RA comparison. In the pre-weighting cohort, GLP-1 RA use was associated with decreased hazard of MACCE compared to bupropion-naltrexone (HR 0.50 [95% confidence interval (CI) 0.36-0.69]) and phentermine-topiramate (HR 0.43 [95% CI 0.30-0.60]). In the propensity score-overlap weighted cohort, GLP-1 RA prescription was not associated with a lower hazard of MACCE than bupropion-naltrexone (aHR 0.69 [95% CI 0.47-1.00]) but was associated with a lower hazard compared to phentermine-topiramate (aHR 0.61 [95% CI 0.41-0.91]; adjusted absolute rate difference 0.98 per 1000 person-years).

**Conclusions:** Prescription of a GLP-1 RA was associated with a lower risk of subsequent MACCE than phentermine-topiramate.

## INTRODUCTION

The Semaglutide and Cardiovascular Outcomes in Obesity without Diabetes (SELECT) trial established the efficacy of semaglutide for the prevention of cardiovascular events among patients with overweight/obesity and at elevated cardiovascular risk but without diabetes mellitus (DM). SELECT led to a United States (US) Food and Drug Administration (FDA) label for this indication,^1^ leading to widespread use of semaglutide in those without DM. However, the magnitude of cardiovascular risk reduction in routine clinical practice, and the comparative effectiveness of glucagon-like peptide-1 receptor agonist (GLP-1 RA) therapy against established second-generation antiobesity agents (including bupropion-naltrexone and phentermine-topiramate) is not known.

Limited evidence exists to compare the effectiveness of these medications, and prior randomized trials of bupropion-naltrexone and phentermine topiramate were halted early and did not definitively establish cardiovascular benefits.^2,3^ Head-to-head trials of these agents are unlikely to be conducted. One retrospective, observational cohort study of 205 patients from South Korea evaluated weight loss between several classes of anti-obesity medications. However, the small size of this study precluded adjustment for confounding factors, and there were limitations in generalizability and a lack of ascertainment of cardiovascular endpoints.^4^ Given the high cost of the latest generation of anti-obesity medications, understanding the comparative effectiveness of these medications is essential.^5^ Therefore, this study aimed to examine the comparative effectiveness of these medications in routine clinical practice.

## METHODS

### Data Sharing Statement

Data can be obtained from Truveta under an approved Data Use Agreement.

### Study Design

This was a retrospective, observational cohort study using a target trial emulation design according to a pre-specified statistical analysis plan (**Supplemental eMaterial).**

### Data Source

Data were obtained from Truveta, which maintains a large, nationwide database of electronic health records from a network of US healthcare systems. The data include structured, harmonized data from sources including patient demographic information, vital signs, laboratory measurements, diagnosis codes from encounters and billing claims, prescription drug requests, procedures, and linked data on social drivers of health (via linkage with LexisNexis).^6^

### Ethical Approval and Reporting

This study was determined to be exempt from institutional review board approval by the Duke University Institutional Review Board (Pro00115475). The study was reported according to the Reporting of Studies Conducted using Observational Routinely-collected health Data (RECORD) statement (**see Supplemental eMaterial).**

### Study Population

Inclusion criteria for the study were 1) age >=18, 2) at least one year of prior data availability, 3) prescription of any of the following: any glucagon-like peptide 1 receptor agonist (GLP-1 RA), the combination of bupropion and naltrexone, the combination of phentermine and topiramate, or orlistat, 4) Prior stroke, transient ischemic attack, myocardial infarction, coronary artery disease, heart failure, or peripheral artery disease, and 5) body mass index (BMI) of 27 kg/m^2^ or higher. Exclusion criteria were 1) history of diabetes mellitus, 2) use of any antidiabetic medication (oral hypoglycemic agent or insulin) other than metformin in the year preceding the index date, or 3) hemoglobin A1c (HbA1C) >6.5% in the year preceding the index date.

### Exposures

The primary exposure was antiobesity-labeled GLP-1 RA prescription, specifically liraglutide, semaglutide, or tirzepatide. The primary comparator was bupropion-naltrexone prescription, given the strongest evidence of cardiovascular benefit for this drug class.^2^ Comparisons with phentermine-topiramate and orlistat were pre-specified as secondary exposures. The index date was determined as the date of the first prescription of either antiobesity medication in each comparison, to concord with the intention-to-treat principle and to minimize the risk of introducing immortal time bias. A schematic for the determination of the index date is shown in the **Supplemental eMaterial.**

### End Points

#### Primary End Point

The primary endpoint was the time to first major adverse cardiovascular/cerebrovascular event (MACCE), defined as stroke or myocardial infarction. A validated algorithm to identify recurrent MACCE events was adapted and classified this endpoint based on the time to first event (e.g., a patient with a history of stroke in the lookback period would be classified as experiencing MACCE at the time they had either a recurrent stroke or an incident myocardial infarction).^7^ In brief, this algorithm integrates both billing, laboratory, and procedure information to discriminate between patients who are admitted for complications of a prior cardiovascular event versus those who have experienced a recurrent event. Precise details of our strategy to ascertain the primary and secondary endpoints are shown in the pre-specified analysis plan (**Supplemental eMaterial).**

#### Secondary Clinical End Points

Secondary clinical end points included all-cause mortality, a composite of recurrent MACCE and hospitalization for heart failure, diagnosis of type 2 diabetes mellitus, diagnosis of chronic kidney disease stages 3-5, and diagnosis of metabolic-associated steatotic liver disease.

#### Secondary Intermediate End Points

Secondary intermediate endpoints included change in BMI, systolic blood pressure, low-density lipoprotein level, high-density lipoprotein level, triglyceride level, and hemoglobin A1c at six months of follow-up.

### Statistical Analysis

#### Covariates

To account for baseline differences between patients prescribed GLP-1 RA versus non-GLP1-RA antiobesity medications, the following covariates were included when deriving propensity scores:

- Demographics (age, sex, race, and ethnicity)
- Clinical comorbidities (all individual components of the Elixhauser comorbidity index and pyelonephritis, pregnancy, osteoarthritis, obstructive sleep apnea, COPD, non-alcoholic fatty liver disease, polycystic ovarian syndrome, coronary artery disease, hyperuricemia, schizophrenia, and traumatic brain injury).
- Baseline laboratory measures (including hemoglobin A1c, LDL cholesterol, HDL cholesterol, total cholesterol, serum triglycerides, serum creatinine, aspartate aminotransferase, alanine aminotransferase, white blood cell count, and hemoglobin)
- Baseline physical measures (including BMI, systolic and diastolic blood pressures)
- Measures of healthcare utilization (number of inpatient, outpatient, and emergency department visits)
- Cardiovascular medication utilization (antihypertensives, antithrombotics, and lipid lowering therapies)
- Social determinants of health data (including number of address changes in the preceding twelve months, time since last recorded eviction, percentile ranking of Lexis Nexis forecasted healthcare costs, and median household income for the neighborhood of the current address).

Several variables indicative of social determinants of health were excluded due to very low completion rates (despite being included in the original analytic plan). Platelet count was also excluded due to high missing rates. Implausible physical measurements and laboratory values were excluded systematically (see details in **Supplemental eMaterial).**

All statistical analysis was performed according to a pre-specified statistical analysis plan which was committed prior to performance of any modeling. Descriptive statistics were presented using counts and percentages for categorical variables and medians with first and third quartiles for continuous variables. Differences in baseline characteristics between the groups were summarized with standardized mean differences (SMD). For all endpoints, overlap weighting was used to balance the characteristics of patients in each arm.^8,9^ For time-to-event end points, Cox proportional hazard models were used to estimate hazard ratios with their corresponding 95% confidence interval (CI), and the proportional hazards assumption was assessed. Patients were censored at the end of data availability or at death (for non-death endpoints). The maximum follow-up duration was set at the time when 85% of the cohort had been censored. For continuous endpoints, the average treatment effect in the treated population (ATT) was estimated, reflecting the difference in means for each endpoint. Missing rates were nearly zero for demographic and medical history variables, but were higher for laboratory variables (ranging from approximately 25% to 60%). We used multiple imputation with chained equations (MICE) to address missing adjustment covariates.^10^ End points were not imputed as they were felt to be missing not at random. The threshold of statistical significance was set at alpha =0.05 for two-sided tests; no adjustment for multiplicity was made, so results for secondary end points should be considered hypothesis-generating. Two deviations were made from the pre-specified analysis plan: first, the number of patients exposed to orlistat was very low, so we did not proceed to further analysis in this population. Second, the recurrent cardiovascular disease classification algorithm yielded very low event rates, and a large fraction of patients were excluded due to a lack of adjudicated claims data, so two sensitivity analyses using distinct definitions of MACCE (any diagnosis in claims and any diagnosis in EHR data, respectively) were conducted.

## RESULTS

### Baseline Characteristics

Derivation of the study population for each analysis is shown in **eFigure 1**. In total, 58,305 patients were included in the primary analysis, of whom 31,343 (median age 60, 67% female) were prescribed a GLP-1 RA and 27,202 (median age 62, 58% female) were prescribed bupropion-naltrexone; 46,710 were included in the secondary analysis, of whom 31,407 (median age 60, 66% female) were prescribed a GLP-1 RA and 15,303 (median age 57, 74% female) were prescribed PT. Characteristics of the participants are shown in **Table 1**; standardized mean differences are shown in the non-weighted population, as overlap weighting guarantees a standardized mean difference of zero.^8^

**Figure 1.**
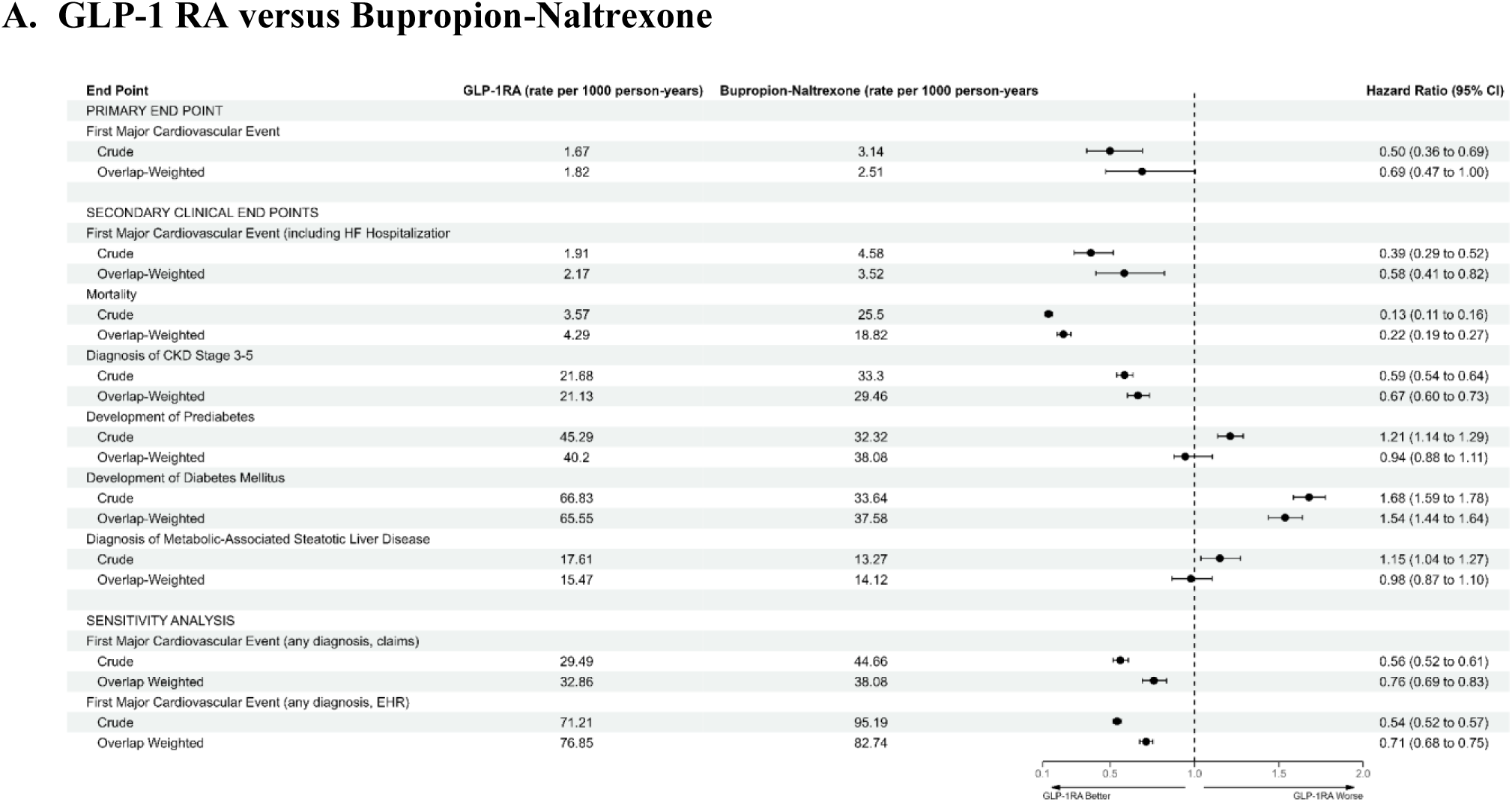
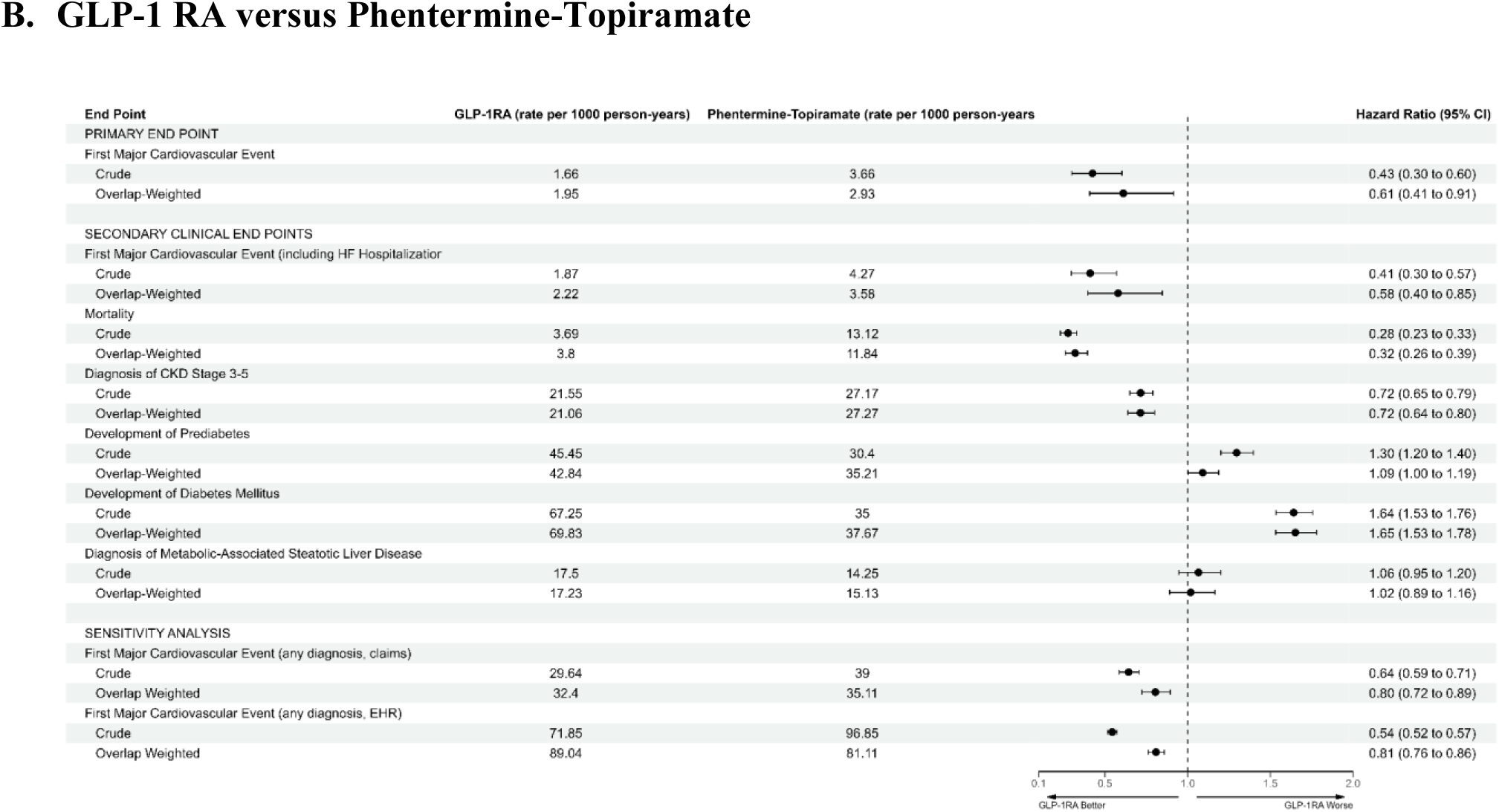
summarizes crude and adjusted rates and hazard ratios for the study endpoints.

**Table 1.** Characteristics of the Study Participants.

| **Characteristic** | **GLP-1 RA versus bupropion-naltrexone** |  |  | **GLP-1 RA versus phentermine-topiramate** |  |  |
| --- | --- | --- | --- | --- | --- | --- |
|  | **GLP-1 RA**<br>**(n=31143)** | **Bupropion**<br>**-**<br>**naltrexone**<br>**(n=27202)** | **SMD** | **GLP-1**<br>**RA**<br>**(n=31407)** | **Phentermine-**<br>**topiramate**<br>**(n=15303)** | **SMD** |
| **Demographics** |  |  |  |  |  |  |
| Age, Median (Q1, Q3) | 60 (52, 68) | 62 (53, 70) | 0.119 | 60 (52, 68) | 57 (47, 66) | -0.24 |
| Sex, No. (%) |  |  |  |  |  |  |
| Female | 20741 (67) | 15892 (58) | -0.17 | 20874 (66) | 11320 (74) | 0.165 |
| Male | 10402 (33) | 11310 (42) | 0.17 | 10533 (34) | 3983 (26) | -0.165 |
| Race, No. (%) |  |  |  |  |  |  |
| White | 24198 (77.7) | 22929 (84.3) | 0.168 | 24463 (77.9) | 11522 (75.3) | -0.061 |
| Black | 4383 (14.1) | 2348 (8.6) | -0.172 | 4382 (14.0) | 2406 (15.7) | 0.498 |
| Asian | 370 (1.2) | 161 (0.6) | -0.064 | 367 (1.1) | 108 (0.7) | -0.048 |
| Other Race | 1940 (6.2) | 1562 (5.7) | -0.021 | 1945 (6.2) | 1107 (7.2) | 0.042 |
| American Indian or Alaska Native | 147 (0.5) | 159 (0.6) | 0.016 | 144 (0.5) | 113 (0.7) | 0.036 |
| Native Hawaiian or Pacific Islander | 105 (0.3) | 43 (0.2) | -0.036 | 106 (0.3) | 47 (0.3) | -0.005 |
| Ethnicity, No. (%) |  |  |  |  |  |  |
| Not Hispanic/Latino | 28128 (90) | 24982 (92) | 0.053 | 28381 | 13644 (89) | -0.04 |
|  |  |  |  | (90) |  |  |
| Hispanic/Latino | 1712 (6) | 945 (3) | -0.098 | 1719 (5) | 977 (6) | 0.039 |
| Other | 1303 (4) | 1275 (5) | 0.024 | 1307 (4) | 682 (4) | 0.015 |
| **Individual Socioeconomic Status** |  |  |  |  |  |  |
| Number of address changes in past 12 months, Median (Q1, Q3) | 0 (0, 0) | 0 (0, 0) | 0.017 | 0 (0, 0) | 0 (0, 0) | 0.093 |
| Time since last recorded eviction, Median (Q1, Q3) | 148 (86, 210) | 145 (83, 219) | -0.021 | 147 (85, 210) | 140 (93, 207) | -0.032 |
| Percentile ranking of Lexis Nexis forecasted healthcare costs, Median (Q1, Q3) | 77 (61, 88) | 82 (67, 92) | 0.175 | 77 (62, 88) | 79 (61, 90) | 0.03 |
| **Community Socioeconomic Status** |  |  |  |  |  |  |
| Median household income for neighborhood of current address, Median (Q1, Q3) | 65560 (47451, 86578) | 61186 (45157, 79276) | -0.178 | 65614 (47480, 86585) | 57698 (43737, 74781) | -0.286 |
| **Laboratory Measures** |  |  |  |  |  |  |
| HbA1c, Median (Q1, Q3) | 5.6 (5.4, 5.9) | 5.6 (5.4, 5.9) | -0.016 | 5.6 (5.4, 5.9) | 5.6 (5.3, 5.8) | -0.113 |
| HDL Cholesterol, Median (Q1, Q3) | 50 (42, 60) | 48 (39, 59) | -0.105 | 50 (42, 60) | 49 (41, 60) | -0.041 |
| LDL Cholesterol, Median (Q1, Q3) | 92 (69.6, 120) | 94 (71, 121) | 0.04 | 92 (69, 120) | 98 (75.0, 124.8) | 0.129 |
| Total Cholesterol, Median (Q1, Q3) | 169 (142, 200) | 170 (143, 200) | 0.018 | 169 (142, 199) | 175 (149, 204) | 0.117 |
| Serum Triglycerides, Median (Q1, Q3) | 115 (85, 159) | 120 (86, 168) | 0.099 | 115 (85, 160) | 117 (84, 165) | 0.057 |
| Serum Creatinine, Median (Q1, Q3) | 0.9 (0.8, 1.0) | 0.9 (0.8, 1.1) | 0.104 | 0.9 (0.8, 1.0) | 0.8 (0.7, 1.0) | -0.023 |
| AST, Median (Q1, Q3) | 21 (17, 26) | 21 (17, 28) | 0.128 | 21 (17, 26) | 20 (16, 26) | 0.041 |
| ALT, Median (Q1, Q3) | 23 (17, 32) | 22 (16, 32) | 0.028 | 23 (17, 32) | 22 (16, 31) | -0.023 |
| White Blood Cell Count, Median (Q1, Q3) | 7.0 (5.8, 8.5) | 7.4 (6.0, 9.2) | 0.214 | 7.0 (5.8, 8.5) | 7.2 (5.8, 8.9) | 0.119 |
| Hemoglobin, Median (Q1, Q3) | 13.7 (12.7, 14.6) | 13.6 (12.4, 14.7) | -0.112 | 13.7 (12.7, 14.6) | 13.3 (12.3, 14.3) | -0.252 |
| Systolic Blood Pressure (nearest to index date), Median (Q1, Q3) | 126 (118, 136) | 126 (118, 138) | 0.03 | 126 (118, 136) | 126 (117, 137) | 0.008 |
| Systolic Blood Pressure (highest in lookback), Median (Q1, Q3) | 146 (134, 162) | 149 (135, 166) | 0.112 | 146 (134, 162) | 148 (134, 164) | 0.067 |
| Diastolic Blood Pressure (nearest to index date), Median (Q1, Q3) | 78 (70, 83) | 76 (69, 82) | -0.106 | 78 (70, 83) | 78 (70, 83) | -0.046 |
| Diastolic Blood Pressure (highest in lookback), Median (Q1, Q3) | 88 (82, 97) | 88 ((81, 99) | 0.061 | 88 (82, 97) | 89 (82, 99) | 0.071 |
| BMI, Median (Q1, Q3) | 37.5 (33.2, 43.1) | 32.6 (29.4, 37.4) | -0.61 | 37.6 (33.2, 43.2) | 34.4 (30.5, 39.7) | -0.379 |
| **Medication Exposures** |  |  |  |  |  |  |
| Angiotensin receptor blocker (ARB), No. (%) | 9651 (31) | 5769 (21) | 0.144 | 9790 (31) | 2789 (18) | 0.318 |
| Angiotensin-converting enzyme (ACE) inhibitor, No. (%) | 6098 (20) | 6757 (25) | 0.102 | 6195 (20) | 3024 (20) | 0.020 |
| Calcium channel blocker, No. (%) | 8399 (27) | 7710 (28) | 0.004 | 8498 (27) | 3758 (25) | 0.001 |
| Beta Blocker, No. (%) | 14593 (47) | 13610 (50) | 0.000 | 14791 (47) | 6861 (45) | 0.002 |
| Potassium-sparing diuretic, No. (%) | 3556 (11) | 2209 (8) | 0.320 | 3615 (12) | 1116 (7) | 0.014 |
| Thiazide diuretic, No. (%) | 8122 (26) | 5644 (21) | 0.029 | 8221 (26) | 3146 (21) | 0.012 |
| Loop diuretic, No. (%) | 6157 (20) | 5305 (20) | 0.111 | 6265 (20) | 2504 (16) | 0.008 |
| Alpha antagonists, No. (%) | 496 (2) | 617 (2) | 0.030 | 509 (2) | 297 (2) | 0.001 |
| Vasodilators, No. (%) | 3865 (12) | 3975 (15) | 0.005 | 3925 (12) | 2245 (15) | 0.013 |
| Anticoagulants, No. (%) | 4240 (14) | 3672 (13) | 0.107 | 4279 (14) | 1709 (11) | 0.002 |
| Antiplatelets, No. (%) | 14313 (46) | 14523 (53) | 0.029 | 14503 (46) | 7523 (49) | 0.031 |
| Statins, No. (%) | 17061 (55) | 15203 (56) | 0.001 | 17275 (55) | 7563 (49) | 0.005 |
| Fibrates, No. (%) | 597 (2) | 580 (2) | 0.005 | 602 (2) | 318 (2) | 0.001 |
| Bile acid sequestrants, No. (%) | 255 (1) | 244 (1) | 0.001 | 259 (1) | 159 (1) | 0.000 |
| Proprotein convertase subtilisin/kexin type 9 (PCSK9) inhibitor, No. (%) | 803 (3) | 251 (1) | 0.717 | 817 (3) | 126 (1) | 0.006 |
| Ezetimibe, No. (%) | 1995 (6) | 1014 (4) | 0.475 | 2024 (6) | 441 (3) | 0.018 |
| Icosapent ethyl, No. (%) | 352 (1) | 362 (1) | 0.000 | 344 (1) | 207 (1) | 0.001 |
| **Healthcare utilization** |  |  |  |  |  |  |
| Number of hospital encounters in prior year, Median (Q1, Q3) | 0 (0, 0) | 0 (0, 1) | 0.172 | 0 (0, 0) | 0 (0, 1) | 0.194 |
| Number of outpatient visits in prior year, Median (Q1, Q3) | 12 (6, 29) | 11 (4, 25) | -0.135 | 13 (6, 29) | 12 (4, 26) | -0.1 |
| Number of emergency department visits in prior year, Median (Q1, Q3) | 0 (0, 1) | 0 (0, 2) | 0.196 | 0 (0, 1) | 0 (0, 2) | 0.297 |
| **Medical History** |  |  |  |  |  |  |
| Hyperlipidemia | 14650 (47) | 9864 (36) | 0.165 | 14788 (47) | 6148 (40) | 0.093 |
| Tobacco Use | 3476 (11) | 4475 (16) | 0.011 | 3550 (11) | 2155 (14) | 0.003 |
| Myocardial Infarction | 2158 (7) | 2192 (8) | 0.000 | 2195 (7) | 1085 (7) | 3.373 |
| Cerebrovascular Disease | 5922 (19) | 5822 (21) | 0.003 | 5913 | 5567 (36) | 0.181 |
|  |  |  |  | (19) |  |  |
| Dementia | 97 (0.3) | 326 (1) | 0.000 | 97 (0.3) | 119 (0.8) | 0.000 |
| Paraplegia and hemiplegia | 253 (0.8) | 276 (1) | 0.000 | 255<br>(0.8) | 349 (2) | 0.000 |
| Hemorrhagic or ischemic stroke | 2884 (9) | 3233 (12) | 0.002 | 2903<br>(9) | 3094 (20) | 0.048 |
| Pyelonephritis | 2219 (7) | 1847 (7) | 0.000 | 2242<br>(7) | 1307 (9) | 0.001 |
| Osteoarthritis | 8719 (28) | 6076 (22) | 0.018 | 8745<br>(28) | 4117 (27) | 0.001 |
| Obstructive sleep apnea | 7220 (23) | 3731 (14) | 0.04 | 7274<br>(23) | 2668 (17) | 0.016 |
| Chronic obstructive pulmonary disease<br>or asthma | 6569 (21) | 5551 (20) | 0.000 | 6589<br>(21) | 3746 (24) | 0.006 |
| NAFLD | 1754 (6) | 679 (2) | 0.003 | 1744<br>(6) | 508 (3) | 0.002 |
| Polycystic ovarian syndrome | 578 (2) | 163 (0.6) | 0.000 | 580 (2) | 192 (1) | 0.000 |
| Coronary Artery Disease | 9127 (29) | 6650 (24) | 0.014 | 9212<br>(29) | 3782 (25) | 0.013 |

In the GLP-1 RA versus bupropion-naltrexone comparison, patients prescribed a GLP-1 RA were older (SMD 0.119), and a greater proportion were female (67% vs. 58%, SMD -0.17) and Black (14.1% vs. 8.6%, SMD -0.175), while a lower proportion were White (84.3% vs. 77.7%, SMD 0.168). Patients prescribed a GLP-1 RA had a lower forecasted percentile of healthcare costs (77th percentile vs. 82^nd^ percentile, SMD 0.175) and a higher neighborhood median household income ($65,560 versus $61,186, SMD -0.178). Laboratory measures were generally similar, as were most medication exposures. For physical measures, patients receiving GLP-1 RA had higher median BMI (37.5 vs. 32.6, SMD -0.61). Medical history variables were similar, although patients receiving GLP-1 RA were more likely to have hyperlipidemia (47% vs. 36%, SMD 0.165).

In the GLP-1 RA vs. phentermine-topiramate comparison, patients receiving GLP-1 RA were younger, less likely to be female, more likely to be White, less likely to be Black, and resided in neighborhoods with higher median household income. Patients in the GLP-1 RA group had lower LDL and total cholesterol at baseline and higher hemoglobin, as well as higher BMI. For medical history, patients in the GLP-1 RA group were more likely to have had a prior myocardial infarction (7.0% vs. 7.1%, SMD 3.373).

### Primary End Point

Crude and adjusted rates of the primary end point (time to first MACCE) are shown in **Figure 1**. Unadjusted cumulative incidence curves for the primary end point are shown in **Figure 2**, and overlap-weighted survival curves are shown in **Figure 3**. Patients prescribed GLP-1 RA had fewer MACCE. Crude and adjusted hazards of the primary endpoint are summarized in **Figure 3**. GLP-1 RA prescription was associated with a lower crude hazard of the primary end point versus bupropion-naltrexone (HR 0.50 [95% CI 0.36-0.69]) and versus phentermine-topiramate (0.43 [95% CI 0.30-0.60]). GLP-1 RA prescription was not associated with a lower adjusted hazard of the primary end point versus bupropion-naltrexone (aHR 0.69, 95% CI 0.47-1.00) but was associated with a lower adjusted hazard of versus phentermine-topiramate (aHR 0.61, 95% CI 0.41-0.91).

**Figure 2.**
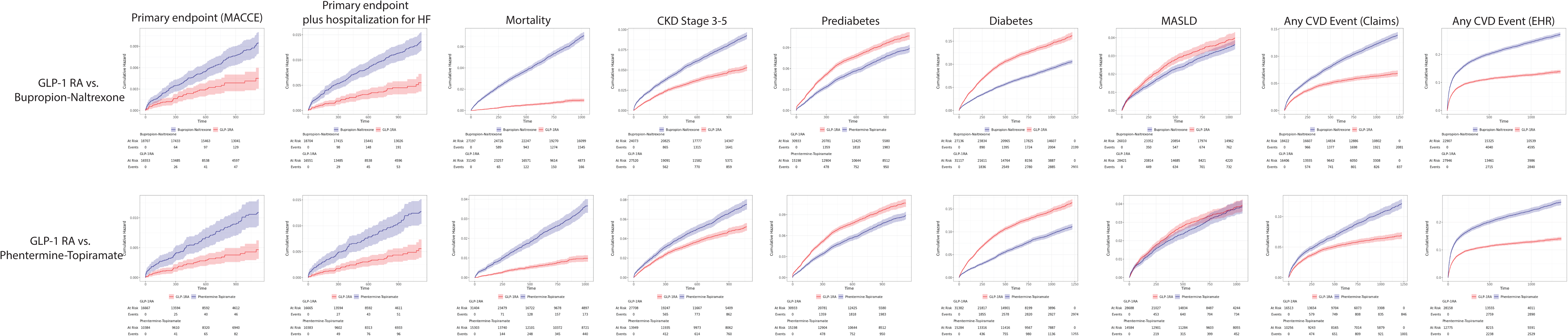
Unadjusted cumulative incidence of the primary and secondary clinical endpoints. **Caption.** Figure 2 shows the unadjusted cumulative incidence of each study endpoint in the two comparisons (GLP-1 RA vs. bupropion-naltrexone and GLP-1 RA vs. phentermine-topiramate, respectively). Overall, rates of all endpoints except diabetes and prediabetes were lower in the GLP-1 RA arm in both comparisons. Survival curves separated early for most endpoints, suggesting the presence of confounding.

**Figure 3.**
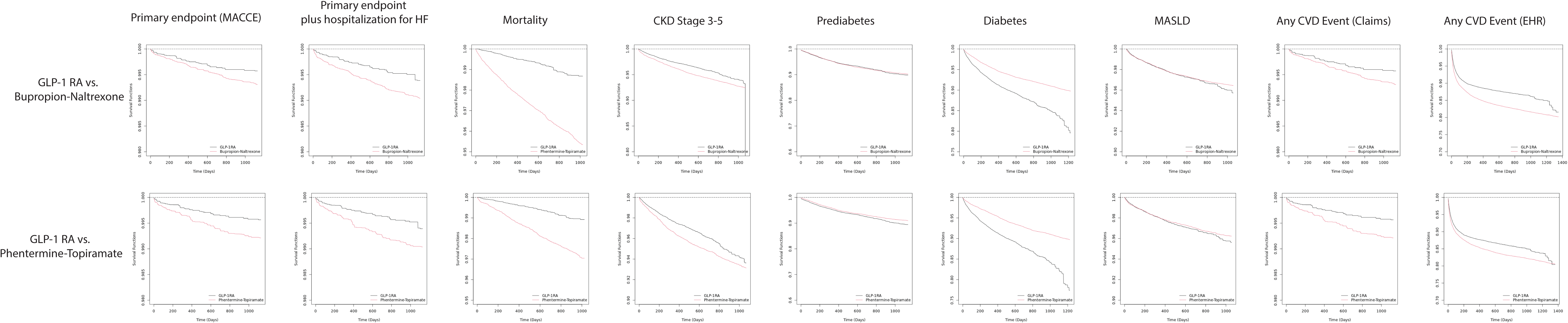
Overlap-weighted survival curves for the primary and secondary clinical endpoints. **Caption**. Figure 3 shows overlap-weighted (adjusted) survival curves accounting for measured baseline differences between patients. Adjusted curves show lower rates of most study endpoints except diabetes diagnosis across the two comparisons, with appropriately delayed separation of survival curves.

### Secondary Clinical End Points

Figure 1 shows crude rates of the secondary clinical end points; Figure 2 shows unadjusted cumulative incidence curves for the secondary clinical end points; and Figure 3 shows overlap-weighted survival curves. In summary, GLP-1 RA prescription was associated with lower crude rates of MACCE (including hospitalization for heart failure), death, diagnosis with chronic kidney disease stages 3-5, and higher crude rates of development of pre-diabetes, type 2 diabetes mellitus, and metabolic-associated steatotic liver disease in both comparison cohorts. After adjustment, as summarized in Figure 3, GLP-1 RA prescription was associated with lower hazards of MACCE (including hospitalization for heart failure), death, and diagnosis of chronic kidney disease stages 3-5. Patients prescribed GLP-1 RA had similar hazards of developing pre-diabetes compared to bupropion-naltrexone but higher hazards of developing pre-diabetes compared to phentermine-topiramate. Patients prescribed GLP-1 RA had higher hazards of Type 2 diabetes compared to both bupropion-naltrexone and phentermine-topiramate, and had similar hazards of diagnosis of MASLD compared to both bupropion-naltrexone and phentermine-topiramate.

### Intermediate End Points

Changes in intermediate end points are shown in **Table 2**. In summary, GLP-1RA prescription was associated with lower BMI, systolic blood pressure, low density lipoprotein cholesterol, high density lipoprotein cholesterol, and serum triglycerides compared to both bupropion-naltrexone and phentermine-topiramate prescription respectively.

**Table 2.** Secondary intermediate endpoints of change in various cardiovascular health indicators for patients taking GLP-1RA versus second-generation antiobesity medications after propensity score overlap weighting.

|  | GLP-1RA versus Bupropion-Naltrexone |  |  |  |  | GLP-1RA versus Phentermine-Topiramate |  |  |  |  |
| --- | --- | --- | --- | --- | --- | --- | --- | --- | --- | --- |
|  | GLP-1RA |  | Bupropion-Naltrexone |  |  | GLP-1RA |  | Phentermine-Topiramate |  |  |
| Endpoint | Baseline (Median, Q1, Q3) | Follow-up (Median, Q1, Q3) | Baseline (Median, Q1, Q3) | Follow-up (Median, Q1, Q3) | Adjusted Absolute Difference* (95% CI) | Baseline (Median, Q1, Q3) | Follow-up (Median, Q1, Q3) | Baseline (Median, Q1, Q3) | Follow-up (Median, Q1, Q3) | Adjusted Absolute Difference* (95% CI) |
| **Body Mass Index (kg/m<sup>2</sup>)** | 37.9 (33.5, 43.4) | 36.6 (32.1, 42.4) | 32.6 (29.4, 37.4) | 32.4 (29.2, 37.2) | -0.94 (-0.94 to -0.94) | 37.9 (33.5, 43.5) | 36.5 (32.1, 42.4) | 34.5 (30.5, 40.0) | 33.8 (29.9, 39.3) | -0.42 (-0.42 to -0.42) |
| **Systolic Blood Pressure (mmHg)** | 126 (118, 136) | 126 (116, 136) | 126 (118, 138) | 126 (118, 138) | -1.43 (-1.43 to -1.43) | 126 (118, 136) | 126 (116, 136) | 126 (118, 137) | 126 (116, 136) | -0.18 (-0.18 to -0.18) |
| **Glycated Hemoglobin (%)** | 5.7 (5.5, 6.0) | 5.6 (5.3, 5.9) | 5.7 (5.5, 6.0) | 5.7 (5.4, 6.0) | -0.13 (-0.13 to -0.13) | 5.7 (5.5, 6.0) | 5.6 (5.3, 5.9) | 5.7 (5.4, 5.9) | 5.7 (5.4, 5.9) | -0.11 (-0.11 to -0.11) |
| **Low Density Lipoprotein Cholesterol (mg/dL)** | 91 (69, 120) | 84 (63, 112) | 92 (70, 119) | 87 (66, 114) | -3.4 (-3.4 to -3.4) | 91 (69, 120) | 84 (63, 112) | 97 (74, 125) | 92 (70, 119) | -4.5 (-4.5 to -4.5) |
| **High Density Lipoprotein Cholesterol (mg/dL)** | 50 (41, 60) | 49 (41, 60) | 48 (39, 59) | 48 (40, 59) | -0.65 (-0.65 to -0.65) | 50 (41, 60) | 49 (41, 60) | 50 (41, 61) | 49 (41, 60) | -0.54 (-0.54 to -0.54) |
| **Serum Triglycerides (mg/dL)** | 118 (87, 162) | 110 (81, 150) | 122 (88, 171) | 119 (85, 168) | -7.9 (-7.9 to -7.9) | 118 (87, 162) | 110 (81, 151) | 117 (84, 162) | 113 (82, 160) | -6.0 (-6.0 to -6.0) |

\* After propensity score overlap weighting

### Post-Hoc Sensitivity Analyses and Contextual Measures

Two alternative definitions of subsequent major CVD events yielded similar results to the primary end point (Figure 3). The E-value for the primary endpoint in the GLP-1 RA vs. phentermine-topiramate comparison was 2.66 for the point estimate and 1.43 for the confidence interval for the phentermine-topiramate comparison.

## DISCUSSION

In patients with a history of cardiovascular disease receiving a GLP-1 RA or second-generation antiobesity medication in routine clinical practice, GLP-1 RA therapies were associated with approximately 20% lower hazard of subsequent major cardiovascular/cerebrovascular events compared to phentermine-topiramate, with no difference in risk compared to bupropion-naltrexone.

The primary result, that there was no difference in cardiovascular event risk between patients prescribed GLP-1 RA and those prescribed bupropion-naltrexone, should be taken cautiously, as the upper bound of the 95% confidence interval was marginally above one (1.004), and event rates were lower than anticipated due to the use of a highly specific algorithm to differentiate recurrent from incident cardiovascular events. Post-hoc sensitivity analyses using broader definitions of this end point showed lower rates of cardiovascular events among patients prescribed GLP-1 RA. Additionally, a pre-specified secondary endpoint adding heart failure hospitalizations to the primary endpoint definition, which increased event rates slightly, estimated a lower hazard of cardiovascular events in the GLP-1 RA arm than the bupropion-naltrexone arm. Taken together, these findings suggest there is likely a decreased risk of cardiovascular events among patients prescribed GLP-1 RA.

Recently, data emerged that supports the promise of medical weight loss therapy to reduce the incidence of MACCE.^11^ The SELECT trial showed an approximately 20% decreased hazard of MACCE for semaglutide 2.4 mg weekly compared to placebo for patients with a history of cardiovascular disease. The present study suggests that GLP-1 RAs may have similar magnitudes of benefit over second-generation and obesity medications in routine clinical practice.^1^ It is possible that GLP-1 RA could be more effective in the prevention of atherosclerotic cardiovascular disease because of greater effectiveness in weight loss or because of pleiotropic effects of GLP-1 RA, including anti-inflammatory effects, improved cardiac function, and hepatic and renal protection, as has been suggested by prior studies.^12–15^ Alternatively, it is also possible that GLP-1 RA therapy might be more effective than second-generation antiobesity medications due to improved safety profile. Phentermine, for example, has been linked with increased blood pressure and heart rate, although these associations are somewhat controversial, and some evidence suggests phentermine-topiramate therapy results in a net reduction of systolic blood pressure.^16–18^

Secondary clinical end points were generally supportive of an improved effectiveness profile of GLP-1 RAs compared to both bupropion-naltrexone and phentermine-topiramate, with lower hazards of chronic kidney disease diagnosis and mortality observed in the GLP-1 RA group.

However, there was an unexpectedly higher rate of diagnosis of prediabetes and type 2 diabetes in the GLP-1 RA group, which is unlikely to be clinically accurate given the evidence that these medications delay or prevent development of these conditions.^19^ Given insurance policies which will often only cover GLP-1 RAs when patients are diagnosed with type 2 diabetes, it is plausible that surveillance bias may drive this association, especially given that our intermediate end point of hemoglobin A1c showed lower six-month A1c measurements among patients prescribed GLP-1 RA versus those prescribed bupropion-naltrexone and phentermine-topiramate.^20^

Analyses of intermediate endpoints such as change in body mass index (BMI), hemoglobin A1c, systolic blood pressure, and serum lipid concentrations were generally supportive of our primary outcome results, as GLP-1 RA prescription was associated with small but consistent reductions in these measures compared to bupropion-naltrexone prescription and phentermine-topiramate prescription. Taken together, these results suggest that improvements in cardiovascular risk factors correlate well with reductions in cardiovascular events, concordant with existing literature and clinical trials.^1,21,22^

This study has limitations. First, the study design compared first time prescription of GLP1-RA versus second-generation antiobesity medications and was analyzed according to the intention-to-treat principle. Therefore, this study does not capture the effects of long-term adherence, which may be different between the two medication classes, and our results accordingly may underestimate the impact of GLP-1 RA with consistent adherence.^23^ Second, to maximize the internal validity of this study, a definition for the primary endpoint that exhibits high specificity (relying on both billing data and laboratory/procedure data) resulted in lower-than-expected event rates, particularly for acute myocardial infarction; however, a post-hoc sensitivity analysis using a definition with potentially greater sensitivity found broadly similar results. Third, this study is susceptible to confounding by indication, although a robust set of adjustment variables were used (including laboratory, physical measurement, medication, and social determinants of health data that is often unavailable in data sources such as claims databases). This likely helped minimize the potential impact of confounding by indication, as suggested by the estimated E-values for the point estimates both populations, which suggested that an unmeasured confounder would need to increase the relative risk of the outcome by nearly threefold, which is unlikely to be the case in the context of the broad adjustment variables already used; additionally, adjusted survival curves appropriately diverge slowly after the index event.^24^ Fourth, this study’s secondary intermediate endpoints (e.g. change in BMI, systolic blood pressure, etc.) should be interpreted with caution, as many patients did not have measurements within the eligible window in routine clinical practice, and this requirement inevitably introduces immortal time bias. The primary analysis, however, is not vulnerable to this limitation.

These results have implications for the secondary prevention of cardiovascular disease in routine clinical practice. First, they suggest that the effectiveness of semaglutide in the secondary prevention of ASCVD could potentially extend to a broader patient population than is currently targeted. The results provide additional supportive evidence that GLP-1 RA therapy is associated with risk reduction in routine clinical practice. Additional investigation into the effectiveness of bupropion-naltrexone and phentermine-topiramate for the secondary prevention of ASCVD is indicated, given unclear clinical trial data, and there may be a subset of the population for whom these drugs may be more efficacious. This is an important policy question given the high cost of GLP-1 RA therapy and widespread population eligibility for this indication, especially because a recent economic evaluation suggested that GLP-1 RA therapy is not cost-effective compared to bupropion-naltrexone and phentermine-topiramate for the treatment of overweight/obesity in the general population.^25,26^

## ACKNOWLEDGEMENTS

The authors report no acknowledgements.

## Disclosure of Interest: All authors have completed the ICMJE uniform disclosure form and declare the following

WG: No relevant disclosures.

MW: No relevant disclosures.

JS: No relevant disclosures.

FL: No relevant disclosures.

EO: No relevant disclosures.

LG: Glover reports grant funding from the Trans-Omics for Precision Medicine, the National Institutes of Health, and the National Heart, Lung, and Blood Institute

KB: No relevant disclosures.

AZ: No relevant disclosures.

RM: McDevitt reports grant funding from Duke University Office of the Provost, Arnold Ventures, Washington Center for Equitable Growth, and the American Investment Council. McDevitt has received consulting and expert witness fees through Charles River Associates as well as speaker fees from Welsh, Carson, Anderson, & Stowe, In Tandem Capital, and Heritage Group, and in the past served on the advisory board of Renalogic.

CK: No relevant disclosures.

SW: No relevant disclosures.

SS: No relevant disclosures. SA: No relevant disclosures.

MP: No relevant disclosures.

BMG: Dr. Mac Grory reports grant funding from the National Institutes of Health (K23HL161426, UG3NS138219, and R03HL178686), the American Heart Association (23MRFSCD1077188 and 25GLP1450119), the Duke Office of Physician Scientist Development (2835124), the Duke Clinical Research Institute, and the Duke University Office of the Provost.

JBL: Dr. Lusk reports grant funding from the American Heart Association, Alzheimer’s Association, National Institute on Ageing, Health Resources and Services Administration, and Duke University Office of the Provost.

## DATA AVAILABILITY STATEMENT

Data can be obtained from Truveta under an approved data use agreement.

